# Expert-validated estimation of diagnostic uncertainty for deep neural networks in diabetic retinopathy detection

**DOI:** 10.1101/19002154

**Authors:** Murat Seçkin Ayhan, Laura Kühlewein, Gulnar Aliyeva, Werner Inhoffen, Focke Ziemssen, Philipp Berens

## Abstract

Deep learning-based systems can achieve a diagnostic performance comparable to physicians in a variety of medical use cases including the diagnosis of diabetic retinopathy. To be useful in clinical practise, it is necessary to have well calibrated measures of the uncertainty with which these systems report their decisions. However, deep neural networks (DNNs) are being often overconfident in their predictions, and are not amenable to a straightforward probabilistic treatment. Here, we describe an intuitive framework based on test-time data augmentation for quantifying the diagnostic uncertainty of a state-of-the-art DNN for diagnosing diabetic retinopathy. We show that the derived measure of uncertainty is well-calibrated and that experienced physicians likewise find cases with uncertain diagnosis difficult to evaluate. This paves the way for an integrated treatment of uncertainty in DNN-based diagnostic systems.

## Introduction

Deep neural networks (DNNs) are emerging as powerful tools for medical image analysis and disease diagnosis (reviewed by^1, 2^). It has been shown that DNNs can detect diabetic retinopathy (DR) from fundus images^3^ or skin cancer from dermoscopic images^4, 5^ with high accuracy. These and similar studies^6–8^ established that DNNs can achieve or even surpass human level performance on challenging diagnostic tasks, raising the hope that deploying DNNs in clinical settings may improve clinical workflows by automating certain tasks. For instance, a CNN-based image mining tool for DR detection has been integrated into a mobile screening system^9^ and first DNN-based systems have been approved by the U.S. Food and Drug Administration (FDA)^10^. With the help of DNNs, the opportunistic screening could be replaced by a widespread systematic approach and carried out directly by family doctors and primary care physicians^11–13^.

Despite this impressive performance, DNNs do not generate well-calibrated, reliable uncertainty estimates regarding their decisions^14–18^. In fact, the deeper the network, the less-well is it typically calibrated^15^. On the other hand, the interobserver variability in human reading of medical images is high^19–23^, which suggests that traditional screening methods can miss significant numbers of disease cases, especially in large cohorts. Considering that human annotations (*labels*) are used to supervise the training of DNNs, it is imperative to obtain reliable uncertainty estimates^24^ so that clinical professionals can properly judge whether an automated diagnosis can be trusted in any given instance.

Ideally, one would formulate DNNs in a Bayesian framework to obtain well-calibrated uncertainty estimates. While this is mathematically straightforward^25–27^, such Bayesian DNNs are computationally intractable for many real world problems^14, 16, 17^. Recently, tractable solutions to this problem have been proposed. For example, it has been shown that using dropout – originally developed as a regularizer – at test time can provide viable uncertainty estimates^14^, also in a medical setting^28^. This method is called Monte Carlo Drop-Out (MCDO). However, state-of-the-art network architectures^29–34^ use Batch Normalization (BN)^35, 36^ as an implicit regularizer^37, 38^ instead of explicit dropout layers. BN has also been cast as an approximate Bayesian inference method called Monte Carlo Batch Normalization (MCBN)^39^ where one uses the stochasticity of minibatch statistics in order to obtain uncertainty estimates^39^. However, the quality of such Bayesian uncertainty estimates strictly depends on the suitability of the prior and the approximation efficacy which is usually tied to computational constraints^17^. Moreover, MCBN trades the network’s maximum classification performance for uncertainty (see in Discussion).

Non-Bayesian alternatives can offer simpler yet effective means to quantify the uncertainties of DNNs. Here, we build on recent ideas^42^ using test-time data augmentation (TTAUG) to estimate the diagnostic uncertainty in DNNs in an intuitive and data-driven way. Briefly, the network is presented with a number of subtle variations of the input images and we record the network’s responses to such changes in the vicinity of examples in the input spaces. From this distribution over network outputs, we can derive an estimate of predictive uncertainty. Importantly, this technique can be applied to state-of-the-art networks regardless of their design choices or regularization methods. We apply TTAUG to the case of detecting DR from fundus images, a well understood diagnostic task, for which the high performance of DNNs has been demonstrated. We find that TTAUG not only yields well-calibrated uncertainty estimates, but also makes the network more robust on previously unseen data. Furthermore, we validate our uncertainty measures clinically, by showing that disagreement between clinicians is higher for images for which the network reported high uncertainty. This is a crucial prerequisite for making such uncertainty estimates useful in practice.

## Methods

### Bayesian Deep Neural Networks for Medical Diagnostics

In a supervised scenario, a DNN is essentially a sophisticated function that maps inputs to outputs: *y* = *f*_*θ*_ (**x**), where θ represents the network’s free parameters. In order to infer an optimal configuration of *θ*, the network is typically trained on a finite dataset 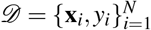. To obtain a probabilistic view on this issue, DNNs can be viewed from a Bayesian perspective:

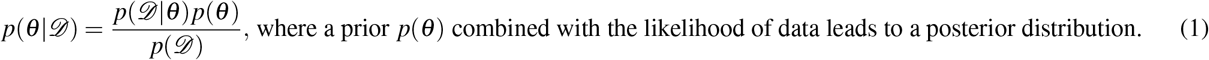

Then, the network can be used for predictions on new examples as follows: 𝔼[*y*] = ∫ *f*_*θ*_ (**x**)*p*(*θ 𝒟*)*dθ*. Despite the elegance of the Bayesian framework, it is intractable to directly work with this integral, as the parameter space for DNNs is extremely high-dimensional. Typically, we do not have access to the true posterior over the network parameters: *p*(*θ* |*𝒟*). Instead, we resort to a single point estimate: *maximum likelihood estimate (MLE)*, which leads us to point predictions.

A DNN achieves classification typically via a *softmax* function in its classification layer and estimates the class probabilities of an image **x** as follows: 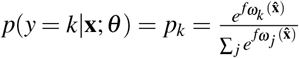, where *k* is an index into *K* classes, *ω*_*k*_ ⊂ *θ* represents the weights and bias for the *k*-th class in the softmax layer, 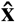 is the feature representation by the network’s penultimate layer, and outputs are multinomial distributions: ∑_*k*_ *p*_*k*_ = 1.

### Datasets and Disease Detection Tasks

We evaluate our method using two collections of fundus images: (i) a data set from a Kaggle competition^43^ and (ii) the Indian Diabetic Retinopathy Image Dataset (IDRiD)^44^. The Kaggle DR dataset consists of 35,126 training images and 53,576 test images graded according to the International Clinical Diabetic Retinopathy Severity Scale^45, 46^: 0-No DR, 1-Mild DR, 2-Moderate DR, 3-Severe DR and 4-Proliferative DR. The dataset is severely imbalanced and it is dominated by the examples of No DR, which corresponds to ∼73% of images in both the training and test sets. The class priors based on the training set are 73.48%, 6.95%, 15.06%, 2.48%, and 2.01%, respectively. During the competition, the labels of 10,906 test images were public and used by participants for validation purposes. The rest was private and internally used for ranking submitted solutions. In our study, we also adhere to the same partitions of data. The IDRiD dataset consists of 516 retinal fundus images acquired via a Kowa VX-10*α* digital fundus camera with 50° field of view and graded w.r.t. the aforementioned DR scale^44^. In general, IDRiD images are of better quality and contain much less camera artifacts than Kaggle images. The class priors are 32.56%, 4.85%, 32.56%, 18.02%, and 12.02% in regards to the DR scale. We use the entire IDRiD data to test the generalization performance of our network and uncertainty measures.

We trained the network to discriminate all five levels of the DR scale above. We evaluate it for two binary disease detection tasks, considering either Mild DR or Moderate DR as the disease onset. To this end, we dichotomize the network outputs by summing up the softmax values accordingly, resulting in the following groupings: {0} vs. {1,2,3,4} and {0,1} vs. {2,3,4}, respectively. In these detection tasks, *p*(*y* = 1|**x**; *θ*), where 1 marks the presence of disease, is sufficient to indicate the most likely decision.

### Deep Neural Network Architecture and Training

We implemented a CNN based on the residual learning with *bottleneck* design^29, 30^ (Fig. 1a, Table 1 in supplements). We modified the original architecture^29^ using an additional fully connected layer before softmax. Also, our network uses parametric ReLUs (PReLUs)^47^ in the first convolutional stack and fully connected layer. Since these parts of the network have no residual connections, PReLUs promote gradient propagation through those layers. The intermediate convolutional stacks adopt the residual architecture with ReLUs. Head nodes use max pooling, if necessary, to downsample their features maps as well as 1 × 1 convolution in order to match the number of channels to that of the corresponding residual parts. Our residual blocks also adopt EraseReLU^48^, which eliminates the last ReLU in each residual block and reduces non-linearity in the hopes of easing optimization and improving the generalization performance. All weight layers in the network use *Batch Renormalization (BReN)*^36^. We concatenate the max and average pooled features from the convolutional stack before the fully connected layer. Finally, 512 features from the penultimate layer are fed into a 5-way softmax for classification.

**Figure 1.**
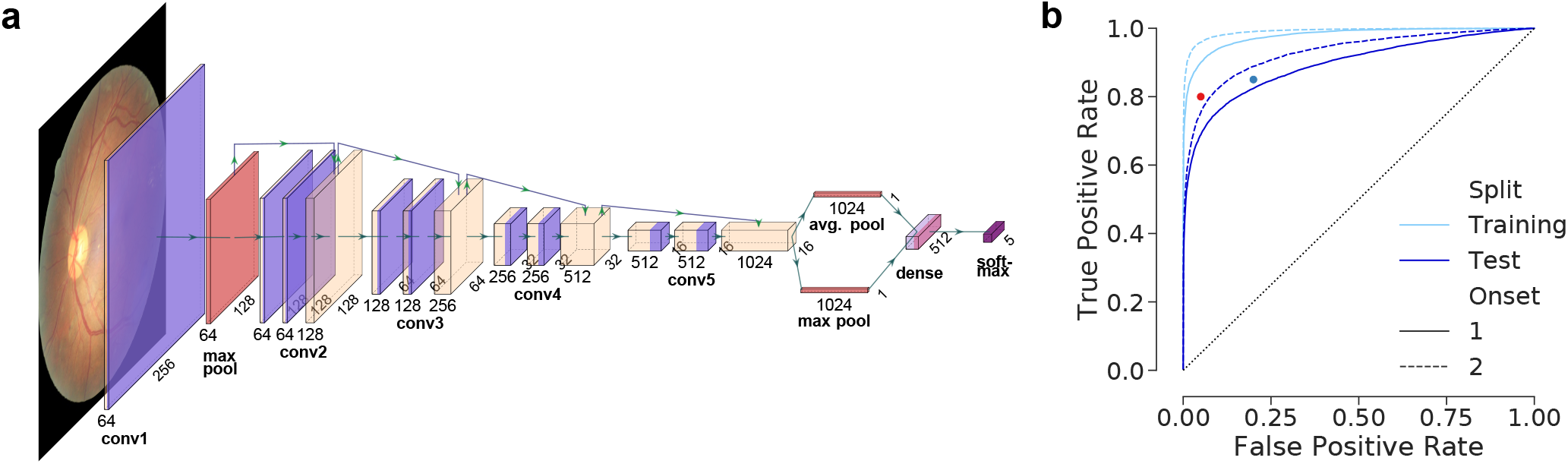
Network architecture and single prediction performance on disease detection tasks. **(a)**: A CNN model with residual connections and 5-way *softmax* function, plotted with PlotNeuralNet^40^. **(b)**: Receiver Operating Characteristic (ROC) curves w.r.t. onset levels 1 and 2. Validation results are excluded for clarity. Thresholds^28, 41^ suggested for the detection of sight-threatening (moderate) DR by the British Diabetic Association (BDA) (80%/95% sensitivity/specificity) and the NHS Diabetes Eye Screening programme (85%/80% sensitivity/specificity) are also given as *red* and *blue* dots, respectively. Sensitivity=TPR, specificity=1-FPR.

We construct a 15-layer network and train it with softmax cross-entropy loss in an *end-to-end* fashion for 500,000 iterations with SGDR^49^ with cosine decay. For the first 50,000 iterations, we use balanced minibatches of 20 images account for severe class imbalance. Then, we increase the minibatch size to 23 and sample stratified minibatches until the end of training, which lets the network adjust itself to the true distribution of classes with an incentive of oversampling^1^. Also note that we apply data augmentation with the probability of 0.9 at each iteration. As a result, the network is exposed to 11,350,000 images, about 10,215,000 of which are randomly generated on the fly.

Weights are initialized via *He’s initialization*^47^ and we use weight decay (*L*2-reg.) with *λ* = 1*e* − 5 for all weight layers except the fully connected layer that adopts *L*1-reg. with the same *λ* to promote the sparsity of features in the penultimate layer. Initial learning rate is 0.003 and it decays with a rate of 0.8 according to the cosine decay schedule, which starts with a period of 10,000 iterations and doubles it after each cycle. The minimum learning rate is set to 1% of the initial rate. The momentum coefficient *µ* is initialized to 0.5 and updated w.r.t. 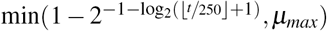^50^, where *t* is the iteration number and *µ*_*max*_ = 0.9, until the 95% of training is completed. Then, we set *µ* = 0.5 once again to allow for finer convergence towards the end of training. The learnable parameters of the BReN layers are initialized as *γ* = 1, *β* = 0 with the exception that the last BReN layers in residual blocks are initialized with *γ* = 0, *β* = 0, which emphasizes the information propagation via the identity connections and eases the optimization, especially in the early stages of training^51^. After all, the performance of the network is validated on the public examples once in 5,000 iterations. The best performing configuration is saved and used for inference. Training curve and model selection based on validation performance are shown in Fig. S1.

We used Tensorflow 1.9-1.13^52^ for the development, training and validation of our networks, all of which were performed on an NVIDIA Titan Xp GPU with 12GB memory and CUDA versions 8/9/9.2 and cuDNN 7. Code and models will be available at https://github.com/berenslab/ttaug-DR-uncertainty upon publication.

#### Image Preprocessing

All images are cropped to a squared center region, resized to 512 × 512 pixels and locally color-normalized for contrast enhancement^28^. For the sake of floating point representation of images, pixel values are mapped from [0, 255] into [0, 1]. Also, all images that are fed to the network are standard normalized w.r.t. the global image statistics.

### Data Augmentation

In addition to image processing, we perform data augmentation before providing images to the network during both training and test times. To this end, we first adopt a recently proposed image acquisition model and its transformation function^53^ *𝒯*, which is parameterized by Φ = [*ϕ*_1_, …, *ϕ*_*n*_] for *n* different transformations: **x** = *𝒯*_Φ_(**x**_0_). In the end, an augmented image **x** is a result of a series of transformations embedded into *𝒯* and applied onto **x**_0_. Assuming prior distributions over the parameters of transformations, the distribution of augmented images *p*(**x**) covers the distribution of observations *p*(**x**_0_)^53^. However, in a classification setting, where the goal is to learn a mapping from images to its assigned class labels, *y* and *y*_0_ are ideally invariant to *𝒯*. In other words, one should refrain from excessive transformations that might severely alter the structure in the data.

We use the following operations in our data augmentation pipeline.

1. Crop and resize: A random variable *crop* controls whether an image will be cropped or not: *crop* ∼ *Bern*(*ϕ*_11_) where *ϕ*_11_ = 0.5. If yes, the lower left and upper right corners of the box to crop into are independently and randomly sampled from the margins of *ϕ*_12_ = 0.15 of height or width from each side: *x*_1_ ∼*U* (0, *ϕ*_12_), *y*_1_ ∼*U* (0, *ϕ*_12_), *x*_2_ ∼*U* (1 −*ϕ*_12_, 1), *y*_2_ ∼ *U* (1 −*ϕ*_12_, 1). Then, the image is resized to its previous dimensions. Note that *ϕ*_1_ = [*ϕ*_11_, *ϕ*_12_].
2. Color transformations are applied w.r.t. factors sampled as follows:
  - Brightness: *b* ∼ *U* (*ϕ*_21_, *ϕ*_22_) where *ϕ*_21_ = −0.15 and *ϕ*_22_ = 0.15
  - Hue: *h* ∼ *U* (*ϕ*_31_, *ϕ*_32_) where *ϕ*_31_ = −0.15 and *ϕ*_32_ = 0.15
  - Saturation: *s* ∼ *U* (*ϕ*_41_, *ϕ*_42_) where *ϕ*_41_ = 0.5 and *ϕ*_42_ = 2.5
  - Contrast: *c* ∼ *U* (*ϕ*_51_, *ϕ*_52_) where *ϕ*_51_ = 0.5 and *ϕ*_52_ = 1.5
3. Geometric transformations include independent horizontal and vertical flips, translation and rotation.
  - Horizontal flip is controlled by *f*_*h*_ ∼ *Bern*(*ϕ*_6_) where *ϕ*_6_ = 0.5
  - Vertical flip is controlled by *f*_*v*_ ∼ *Bern*(*ϕ*_7_) where *ϕ*_7_ = 0.5
  - Translation offsets by pixels in 2D: *t*_*x*_ ∼ *U* (*ϕ*_81_, *ϕ*_82_) and *t*_*y*_ ∼ *U* (*ϕ*_81_, *ϕ*_82_) where *ϕ*_81_ = −25 and *ϕ*_82_ = 25
  - Rotation angle: *r* ∼ *U* (*ϕ*_91_, *ϕ*_92_) where *ϕ*_91_ = −15 and *ϕ*_92_ = 15

### Assessing Calibration

Ideally, the probabilistic outputs of a classifier reflects the true probability of its predictions being correct, its accuracy. Such a classifier is said to be well-calibrated and its outputs can be readily interpreted as confidence scores. However, DNNs are notoriously *miscalibrated* and *overconfident* about their predictions^15, 54, 55^ (Fig. 2b). The concordance between confidence and accuracy can be visualized via reliability diagrams^15, 54–56^, where a classifier is better calibrated as it gets closer to the diagonal.

**Figure 2.**
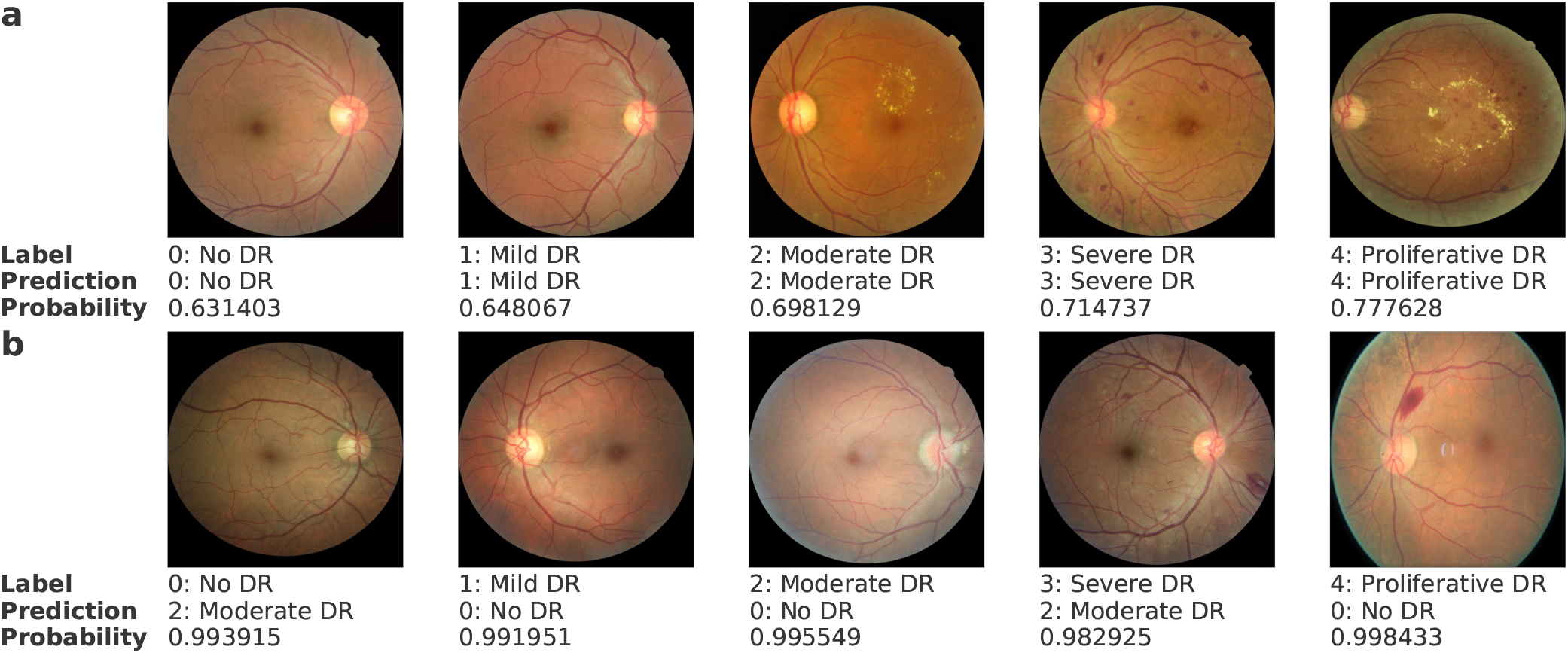
Examples of fundus images with expert labels, network’s predictions and predictive probabilities from the softmax layer. Top row shows correct predictions, whereas overconfident but wrong predictions are at the bottom.

*Expected Calibration Error (ECE)* (Eq. 2) measures the degree of miscalibration, given *M* predictions grouped into *G* bins.

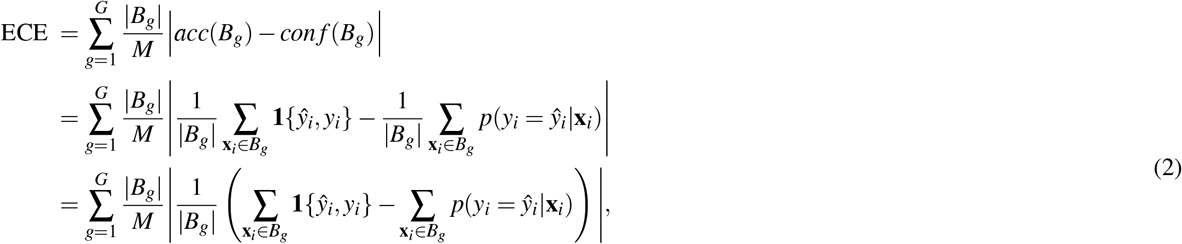

where *ŷ*_*i*_ and *y*_*i*_ are the predicted and actual class labels, respectively. Also, |*B*_*g*_| is the *g*-th bin size, whereas large bars. e.g., | … |, indicate absolute values.

Due to overconfidence, the distribution of predictive probabilities can be highly *non-uniform*. Thus, reliability diagrams and ECE are strongly tied to the binning decisions^55^. While large bins may cause the bin errors to be small, a high-resolution histogram with many bins leads to inaccurate estimation of ECE^55^. Moreover, the gap between the accuracy and confidence of individual predictions may have different signs, which may cause internal compensation inside bins even with uniform distributions^55^. To avoid such complications, we adopt the Adaptive ECE method^55^ that relies on adaptive histograms and computes *positive* and *negative* gaps explicitly.

#### Temperature scaling

Temperature scaling is a parametric approach to calibrating the probabilistic outputs. The basic idea is to rescale the functional values before applying the softmax function so that the probability scores are *softened* and hence better calibrated^15, 57^. Only one parameter, *τ*, needs to be chosen:

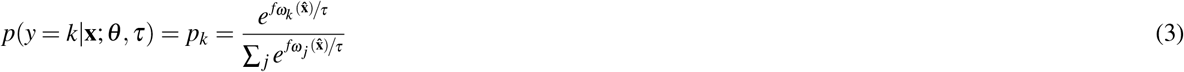

We fit *τ* to the validation data minimizing the negative log-likelihood^15^. Note that *τ* is not an essential part of our network. We use it only to adjust the functional values of the network in a post-processing fashion, which does not change the final decisions. Thus, the accuracy remains untouched, while the probabilities are calibrated.

### Uncertainty measures

Although data augmentation for training is well-established and has been used to improve the discriminative performance of models during inference, its use in predictive uncertainty estimation is underexplored (but see ref^42, 53^). In a probabilistic classification setting, where the classifier outputs are estimated class membership values and only suggest likely class assignments, rather than making deterministic predictions, *y* becomes a random response to examples from *p*(**x**). However, due to a mixture of various types transformations, *p*(Φ) can be a sophisticated joint distribution. Therefore, it is difficult to exactly compute the following quantity to arrive at an aggregate decision with regards to the variation in inputs:

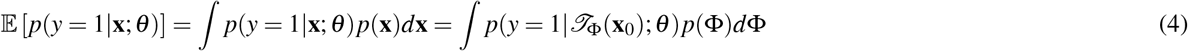

Therefore, we resort to sampling and propose to use TTAUG as an approximate method to evaluate the predictive uncertainty associated with observations. Simply, we sample Φ_*t*_, where *t* ∈ {1, …, *T*}, and generate *T* variations (**x**_*t*_) of a given observation **x**_0_ (Fig. 3a). As a result, we obtain a distribution of predictive probabilities as a proxy for the true but sophisticated one. Then, we can examine how much the network output varies in the vicinity of examples in the input spaces.

**Figure 3.**
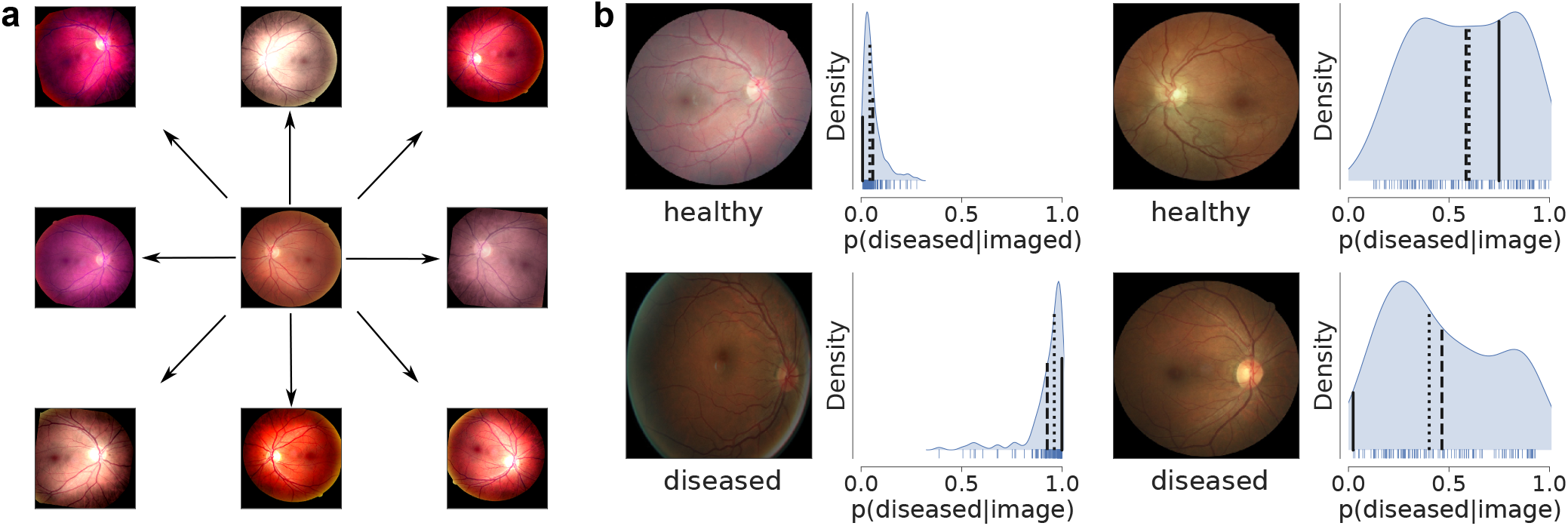
Test-time data augmentation and uncertainty. **(a)**: Given an image in the center, *T* variations are randomly generated via a series of geometric and color transformations. This can be understood in analogy to doctors looking at images from different angles and/or under various illumination so that an underlying disease pattern can be examined from multiple views. **(b)**: Exemplary fundus images (top: healthy, bottom: diseased) and corresponding distributions of predictive probabilities. Given *T* = 128 predictions under onset 1 scenario, mean and median predictions are shown as dashed and dotted lines, respectively. Solid lines indicate the single point predictions. Label assignment is achieved by thresholding the predictions at 0.5. Above the threshold, the label is `diseased`, otherwise `healthy`. Note that the predictive distribution entropy (or spread) is higher in the cases of wrong predictions.

The measures of spread, e.g., standard deviation (STD), interquartile range (IQR), are sufficient under binary classification scenarios. However, these are *undefined* for single point predictions, by which we establish our baseline^58^ performances. Therefore, we use *entropy* (Eq. 5) which is universally applicable to probability distributions.

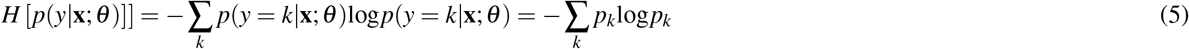

For a binary task, it reduces to *H* [*p*(*y*|**x**; *θ*)] = −*p*log*p*− (1 − *p*)log(1 − *p*), where *p* = *p*(*y* = 1|**x**; *θ*). Given an *ensemble* of *T* predictions 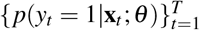, the entropy of the expected prediction *H* [𝔼 [*p*(*y* **x**; *θ*)]] indicates the predictive uncertainty associated with the observation **x**_0_^18, 59^. Typically, 𝔼 [*p*(*y*|**x**; *θ*)] is estimated by a simple mean:

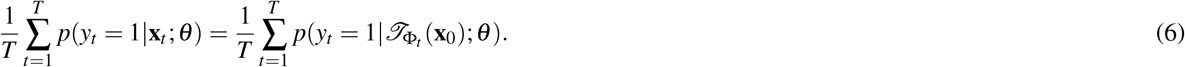

However, *p*(*y* = 1|**x**; *θ*) does not necessarily follow a Gaussian distribution (Figure 3b). Therefore, we additionally test the median prediction for **x**_0_.

### t-distributed Stochastic Neighbor Embedding (tSNE)

tSNE^60^ is a non-linear dimensionality reduction method that enables the interpretation of high-dimensional data in low-dimensional settings. Its main goal is to capture the similarities of examples, ideally based on their high-dimensional feature representations^60^, and map them to a low-dimensional space while preserving the structure in data. In other words, examples that are close/distant in the high-dimensional space must be also close/distant on the low-dimensional map. Probably the most important tSNE parameter is *perplexity*, which effectively determines the size of neighborhood where examples interact with each other. Typical values range from 5 to 50^60^; however, such values may be relatively small for large datasets, especially when one aims at preserving the global structure in data^61^. To this end, we follow the approach of Kobak et al.^61^ and adopt large perplexity values, e.g., ∼ *N/*50, where *N* is the number of examples. Since this quickly becomes computationally infeasible for large *N*, we use Fast Fourier Transform-accelerated Interpolation-based tSNE (FI-tSNE)^62^. Also note that we use the PCA initialization^61^, not only because it promotes the preservation of global structure but also for the reproducibility of tSNE outputs.

### Expert validation

In order to validate our uncertainty estimates, we select a subset of test images and have them re-evaluated by four ophthalmologists according to the same DR grades. One ophthalmologist (GA) was in training with 3 years of clinical experience. The other three are fully trained ophthalmologists or retina specialists with 6-10 years of clinical experience in medical retina. The subset consists of 65 images selected based on the predictive uncertainty under the Onset 1 scenario as well as heuristics for colorfulness^63^ and brightness for the sake of quality in visual inspection. Given the distribution of uncertainty among `correct` and `wrong` predictions (see Fig. S5 in supplements), 5 images are sampled from the following categories: i) wrong and uncertain: 0-No DR, 1-Mild DR, 2-Moderate DR, ii) wrong and confident: 2-Moderate DR, 3-Severe DR and 4-Proliferative DR, iii) correct and uncertain: 0-No DR, 1-Mild DR and 2-Moderate DR, iv) correct and confident: 0-No DR, 1-Mild DR, 2-Moderate DR and 3-Severe DR. We group the images into two w.r.t. predictive uncertainty, which results in low and high uncertainty examples. Then, we compare the DNN decisions with experts’ as well as expert decisions with each other for their coherence. To this end, we use Cohen’s kappa score^64^ that measures agreement between two annotators. Its range is [−1, 1], where 1 indicates a full agreement, whereas lower scores mean less agreement. A negative score can also be interpreted as the degree of disagreement.

## Results

We developed a DNN based on the ResNet architecture to detect diabetic retinopathy from fundus images (Fig. 1a, see Methods). The network was trained with five different categories (healthy and four disease stages), but evaluated only in a binary task, where we considered either mild (level 1) or moderate (level 2) DR as disease onset. The prediction performance of our network was competitive with the recommendations of the British Diabetic Association (BDA) and the NHS Diabetes Eye Screening programme (Fig. 1b), especially given that the recommendations are for the sight-threatening (moderate) DR^28, 41^. However, like many state-of-the-art networks^15^, it provided miscalibrated reports of its uncertainty based on the softmax output (see Methods): While it can correctly classify many fundus images with reasonably high confidence (Fig. 2, top row), it also creates wrong predictions with very high confidence (Fig. 2, bottom row).

To obtain well calibrated predictive probabilities, we generated *T* variations of a given example via subtle modifications (Fig. 3a) and used our network to make a prediction for each augmented example, a procedure we call test-time data augementation (TTAUG)^42^. This resulted in a distribution of predictive probabilities (Fig. 3b). For two examples of correct predictions, this distribution was narrow, indicating that all augmentation examples were classified similarly (Fig. 3b, left). For two examples of incorrect predictions, it was broad and even multimodal, suggesting that the augmentation operations lead to widely varying outputs (Fig. 3b, right). We computed the mean or the median of this distribution as a smoothed estimate of the predictive probability.

Interestingly, these smoothed predictive probabilities obtained through TTAUG closed the generalization gap between training and test results and slightly improved the discrimination performance of the network on the unseen test data compared to the single predictions (Fig. 4a), even with small values of *T* (Fig. 4b). We found that increasing *T* up to 128 continued to increase performance. Of course, there is a trade-off between the efficiency of creating predictions with high *T* and the slight increases in prediction performance.

**Figure 4.**
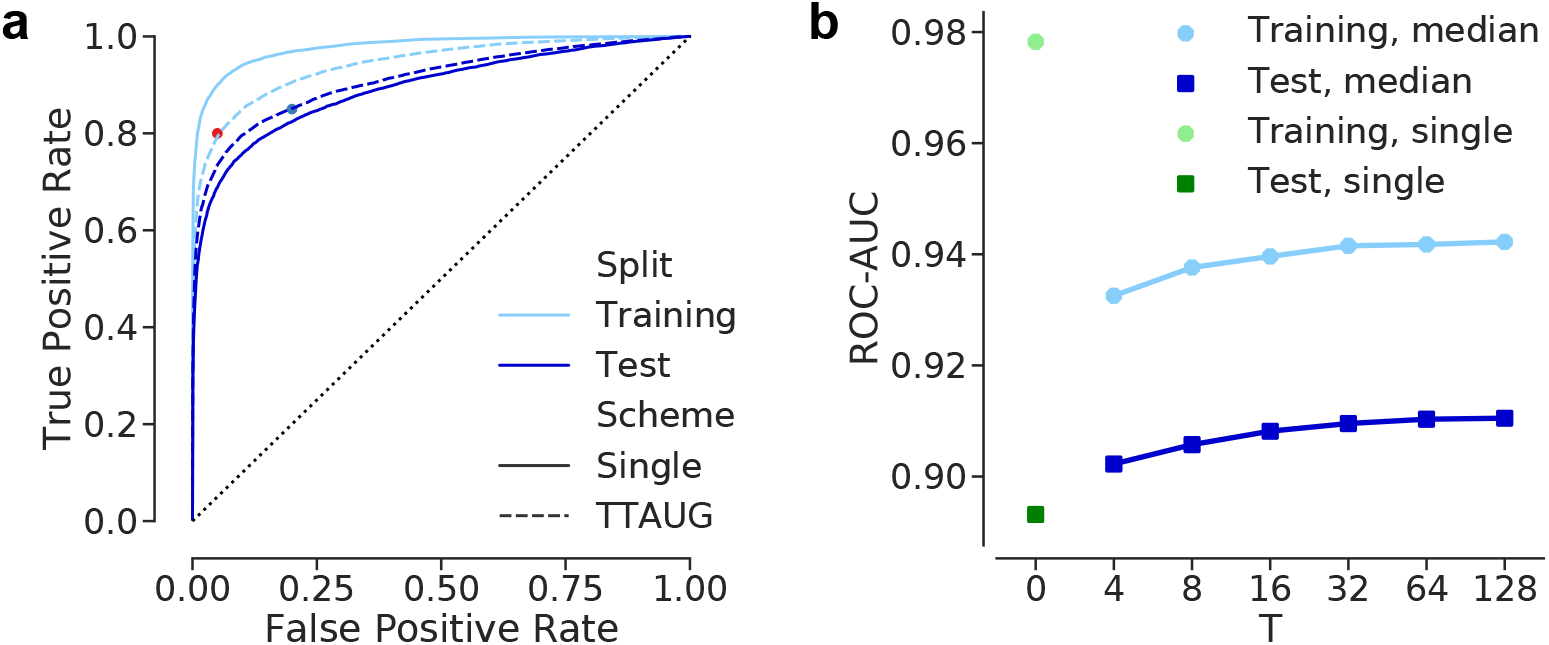
The impact of TTAUG on disease detection performance under onset 1 scenario. Validation results are excluded for clarity. Onset 2 results are given in Fig. S2 in supplements. **(a)**: Receiver Operating Characteristic (ROC) curves for the binary classification via the single and *median* of *T* = 128 predictions. **(b)**: ROC-AUC scores of the single (*T* = 0) and median predictions w.r.t. various *T* values.

We next wondered whether the TTAUG-smoothed estimates of the predictive probability was additionally better calibrated (Fig. 5). Even though our network consisted of only 15 layers, using the single predictive probability led to overconfident predictions with the largest calibration error, as suggested by the examples shown previously (Fig. 2b, Fig. 5a). In the literature, temperature scaling has been suggested to correct for this overconfidence (see Methods). Also in our case, applying temperature scaling led to better calibrated predictive probabilities (Fig. 5b). While the mean of the TTAUG-distribution of predictive probabilities performed worse than the temperature-scaled single predictions with highly underconfident predictions at higher accuracies (Fig. 5c), the median of the TTAUG-distribution of predictive probabilities led to the best overall calibration, both visually and quantitatively (Fig. 5d).

**Figure 5.**
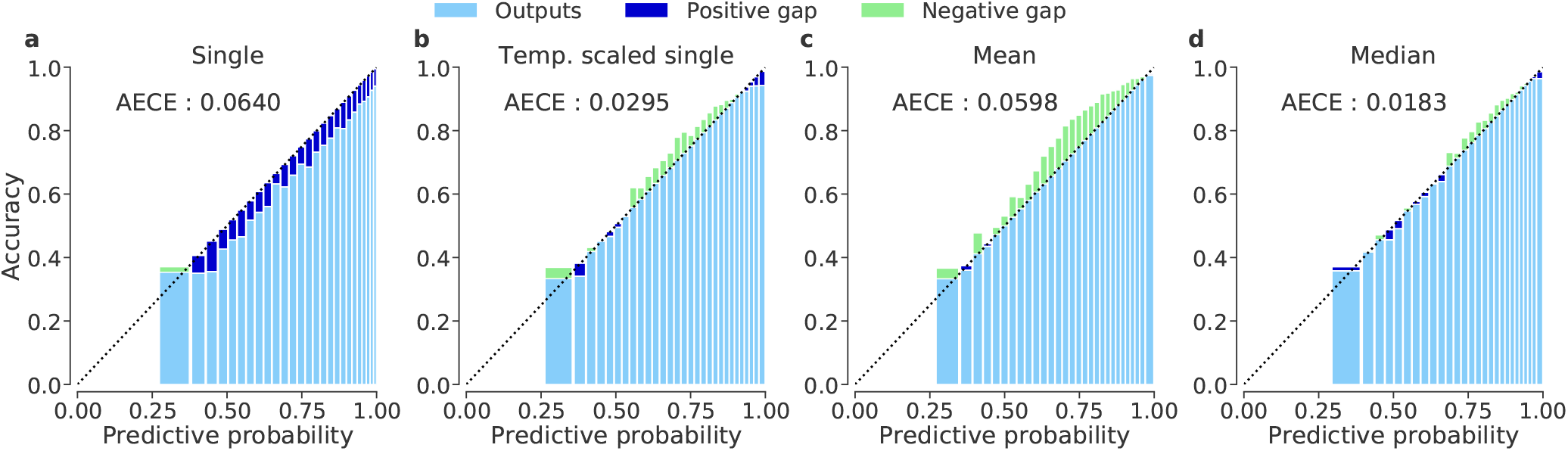
Reliability diagrams and calibration quality via adaptive ECE (AECE) on test data. Positive gap (blue) indicates overconfidence, whereas negative gap (green) implies the lack of confidence. The mean or median of *T* softmax outputs are first computed class channel-wise and then renormalized so that the outputs constitute valid probability distributions. **(a)**: Single prediction. **(b)**: Temperature-scaled single prediction (*τ* = 1.4828538). **(c)**: Mean prediction. **(d)**: Median prediction.

As the median of the TTAUG-smoothed predictive probabilities was calibrated the best, we computed its entropy to quantify the uncertainty of the diagnostic decision of the network (see Methods). To study the properties of this measure, we embed all test examples into one dimension using t-distributed stochastic nearest neighbour embedding (tSNE) based on the network activations in the penultimate layer (Fig. 6a, see Methods). Although the network was presented only categorical labels and has no explicit knowledge of the underlying disease pathology or progression, the map indicates that the network successfully reconstructed the disease continuum and ordered the classes accordingly (compare to a similar 3D tSNE map in^65^) with healthy cases being placed to the left and diseased cases to the right. The cluster of 2064 examples mapped to the leftmost (tSNE < −15) consists essentially of examples with poor image quality, noise or missing content despite the assigned labels (see Fig. S6 for example images from this cluster).

**Figure 6.**
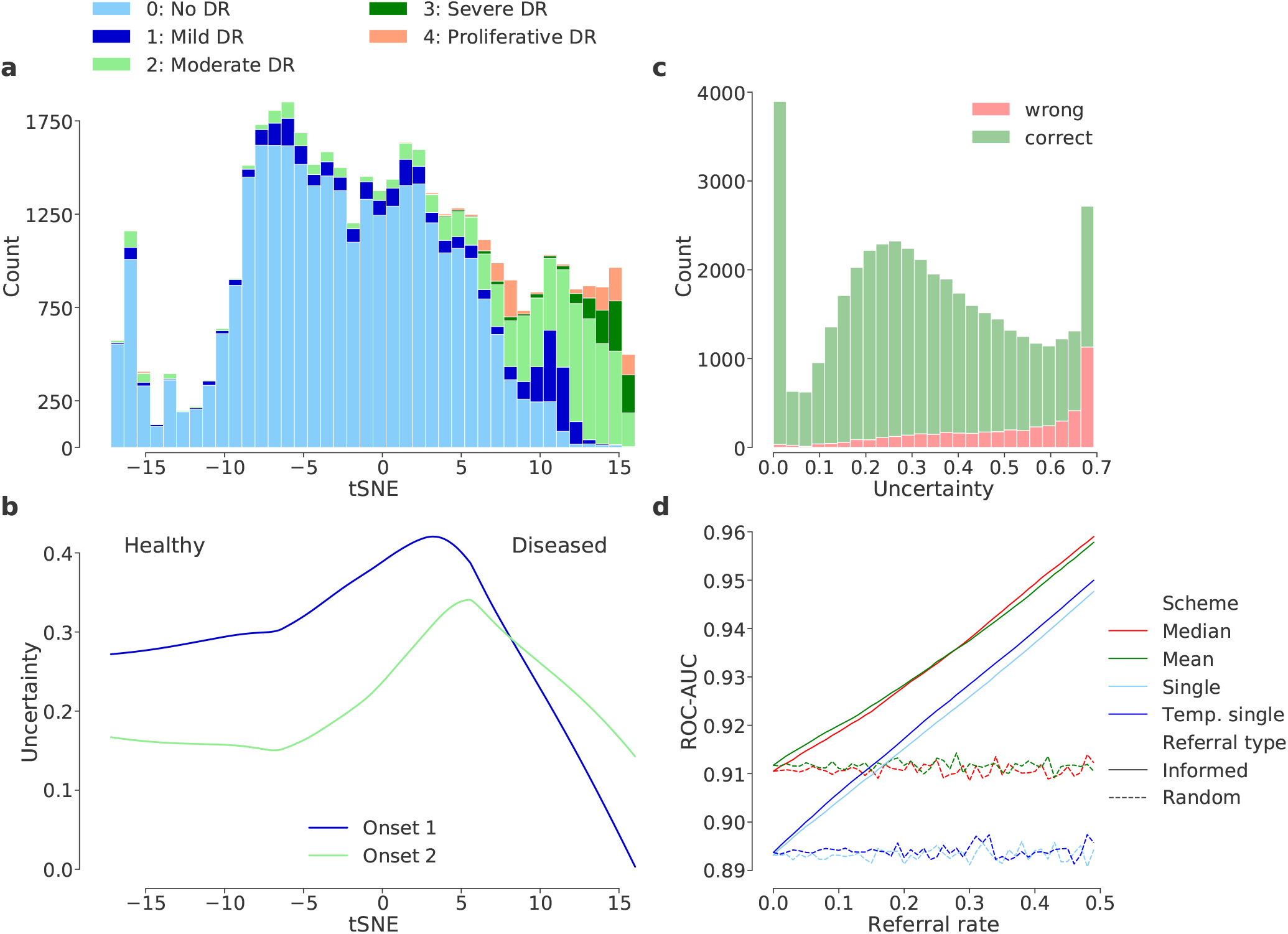
Interpretation of the network’s feature representation and predictive uncertainty for diagnosis, given the test examples. See Fig.S3 in supplements for the onset 2 counterparts of (c) and (d). **(a)** 1D tSNE map via a perplexity of 1000 and 512 features from the penultimate layer. **(b)** Entropy of the median prediction along the disease continuum. Each image is associated with two uncertainty measures under two disease detection scenarios. We fit locally-weighted regression curves to summarize the relations between the tSNE coordinates and predictive uncertainty of examples. **(c)** Distributions of median prediction entropy for all test images grouped by classification results w.r.t the median predictions under onset 1 scenario. **(d)** Improvement in performance via uncertainty-informed decision referral, given different measures of uncertainty under onset 1 scenario. For additional referral plots w.r.t. uncertainty measures derived from the entropy of the entire predictive distribution, see Fig.S4 in supplements.

We found that the average uncertainty was largest at the boundary between disease stages (Fig. 6b). For example, onset 1 uncertainty was highest around t-SNE coordinate 4, where the transition between no DR/mild and moderate DR and the remaining classes could be found. In addition, samples with higher uncertainty were more likely to be erroneously classified (Fig. 6c). This suggests to use the uncertainty measure for *selective prediction*^66–68^, referring predictions on difficult cases for further evaluation by an expert^28^. To compare different uncertainty measures with respect to their ability to refer erroneous predictions efficiently^28^, we ranked the predictions by their uncertainty and measured the network’s performance on the remaining cases, given a certain referral rate. This procedure yielded monotonic improvements in prediction performance, with the median of the TTAUG-distribution of predictive probabilities resulting in the most efficient referrals (Fig. 6d). With this, the network achieved a ROC-AUC score of 0.959 among the remaining data, when 50% of the cases were referred, compared to 0.940 for MCDO at the same referral rate^28^. Our network can achieve this performance at the referral rate of only 32%.

We next evaluated the generalization performance of our network and uncertainty measure on the independent IDRiD data set (see Methods). The network performed even better on the previously unseen IDRiD images (Fig. 7), where it achieved a ROC-AUC score of 0.975 with single predictions under onset 1 scenario. TTAUG with T=128 increased the performance up to 0.982. The improved performance on the IDRiD data is likely due to the quality differences between the respective datasets with less low quality images. While the calibration of network outputs was not as good as it was with the Kaggle DR data (Fig. S7), uncertainty estimates using the entropy of the median of the TTAUG-distribution of predictive probabilities was sufficient to detect perilous predictions (Fig. 7a). As the correct predictions on the IDRiD images had mostly low uncertainty and the incorrect ones mostly high uncertainty, decision referral allows to yield almost perfect classification performance with only 40% of the data referred (Fig. 7b). Interestingly, the uncertainty computed from the temperature scaled single predictive probability fitted to the Kaggle DR validation data did not generalize well to the IDRiD images.

**Figure 7.**
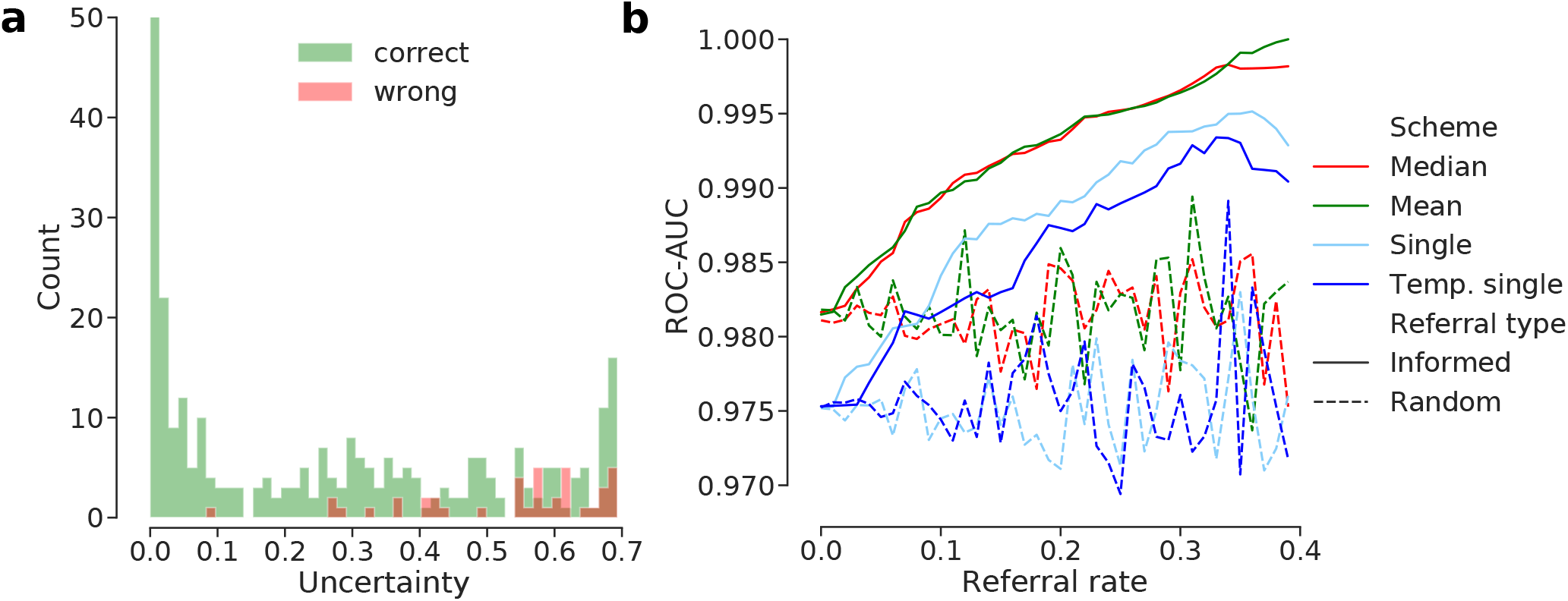
Generalization of the predictive performance and uncertainty estimates to the unseen IDRiD data under the onset 1 scenario. See Fig.S8 in supplements for onset 2. **(a)** Distributions of median prediction entropy for all images from the whole IDRiD data grouped by classification results w.r.t the median predictions. Counts are clipped at 50 for better visualization of the small bins. **(b)** Decision referral on IDRiD images.

Finally, we evaluated whether images with high uncertainty were also more difficult for human experts to grade. To this end, we measured the agreement between the assigned labels and four ophthalmologists as well as among themselves (Fig. 8). First, we selected 65 images covering a range of uncertainties (see Methods and Fig. S5 in supplements), which we divided into two groups of low and high uncertainty based on the entropies of single, mean or median predictions. All images were graded by the ophthalmologists and their gradings were compared to the labels in the data set (for disease onset 1).

**Figure 8.**
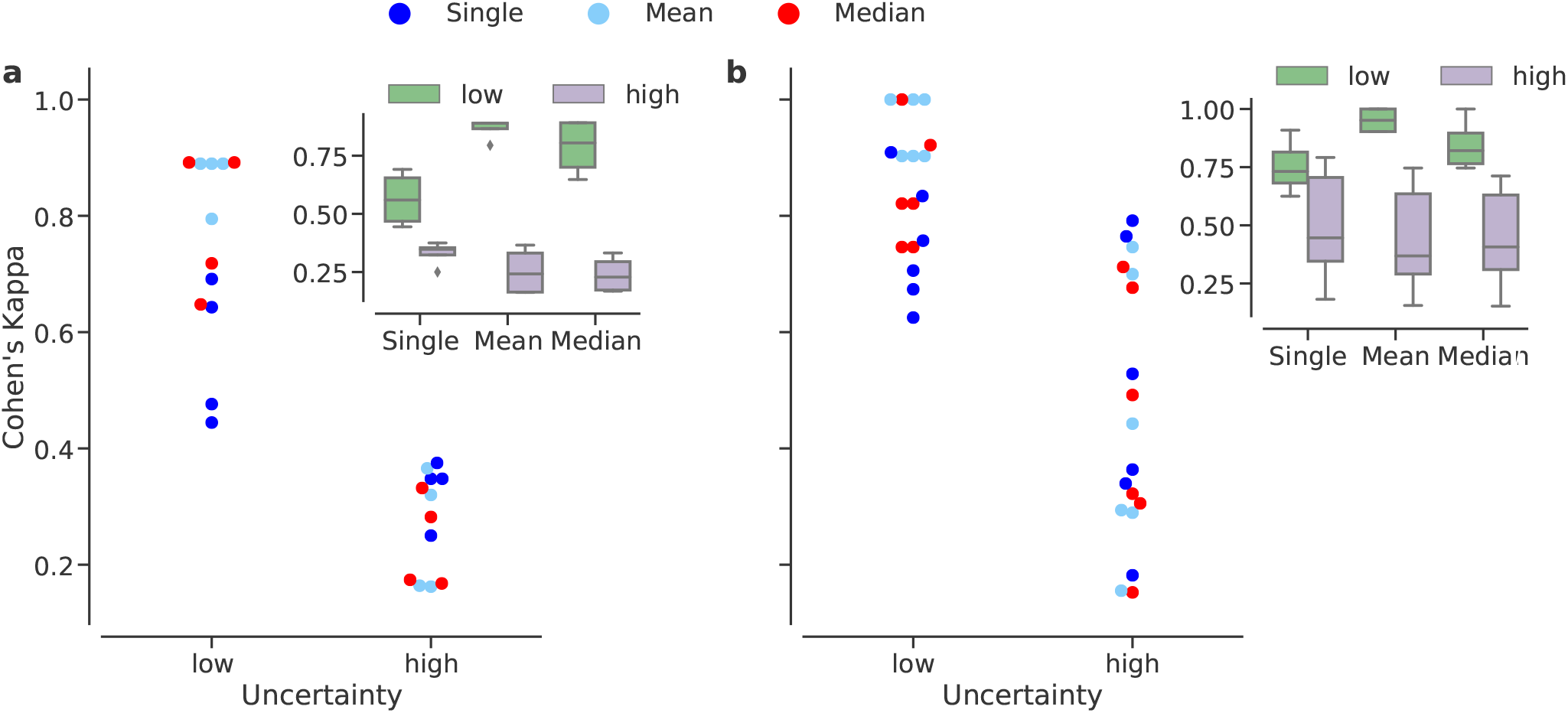
Agreement between annotators under low or high uncertainty from the onset 1 scenario, given Kaggle DR data. Uncertainty groups are determined w.r.t. the predictive uncertainty due to single (blue), mean (light blue) or median (red) predictions. Insets show the box plots for each group within each prediction type. **(a)** The agreement between assigned labels and our expert’s decisions. **(b)** The agreement between experts themselves.

Interestingly, we found that the agreement of judgements by the human experts and the labels in the data set was higher for the low uncertainty images, with a difference in Cohen’s d of .23 ± 0.05, .61 ± 0.05 and .55 ± 0.05 (mean estimate based on ANOVA with SE) between low and high uncertainty cases for single, mean and median uncertainty measures, respectively (Table 2 and 3 in supplements, Fig. 8a). In addition, the agreement was significantly higher in the low uncertainty condition for the mean and median of the TTAUG-distribution of predictive probabilities than the single predictions (ANOVA, post-hoc test with Tukey correction of multiple comparisons, *p* = 0.0003 and *p* = 0.0018, respectively; Table 4 in supplements). Furthermore, we measured the agreement between the four ophthalmologists (Fig. 8b) and found that the inter-annotator agreement on the low uncertainty cases was high, whereas agreement between experts was lower when the uncertainty was high with a difference in Cohen’s d of .27 ± 0.09, .61 ± 0.05 and .55 ± 0.05 (Table 5 and 6 in supplements) for the three measures respectively. Again, the agreement between experts in the low uncertainty condition was significantly higher for the mean and the median of the TTAUG-distribution than for the single predictive entropy (ANOVA, post-hoc test with Tukey correction of multiple comparisons, *p* < 0.0001 and *p* = 0.0019, respectively; Table 7 in supplements). These findings suggest that the TTAUG-based uncertainty measure derived here not only provides better calibration and more efficient referral, but reflects also better the inherent difficulties of trained ophthalmologists in grading a set of fundus images.

## Discussion

Here, we use a state-of-the-art DNN for detecting DR from a well-known dataset with fundus images and evaluate a simple technique to quantify the predictive uncertainty of DNNs. We demonstrate its applicability and benefits for this disease detection scenario with a *human-in-the-loop* mindset. Our network meets the requirements^28, 41^ set forth for the detection of sight-threatening DR (Fig. 1 and Fig. 4) and our uncertainty measures (Fig. 3) are useful for inferring the DR cases that are difficult for the network to classify (Fig. 6). Furthermore, we show that images with high uncertainty are also difficult to grade for physicians with high disagreement between them. This indicates that these images are genuinely hard to classify, a crucial prerequisite for making such uncertainty estimates useful in practice (Fig. 8). The fact that our measures generalize well (Fig.7) to new data also highlights their utility.

Our intuitive and data-driven approach yields a general purpose solution that delivers well-calibrated and accurate uncertainty estimates without requiring substantial changes to the standard training or inference pipelines. In fact, data augmentation is frequently used in training of diagnostic DNNs. Given the variety of transformation operations readily available in many deep learning frameworks or via third party packages, one can flexibly design new data augmentation pipelines and tailor them to the need of new domains and tasks, regardless of the architectural design choices or regularization methods.

### Related Work

The predictive uncertainty of DNNs can be mainly decomposed into two parts: *epistemic uncertainty* and *aleatoric uncertainty*^16, 18^. Epistemic uncertainty can be formalized by means of a probability distribution over the model parameters and accounts for our ignorance about them. It is also known as model uncertainty and can be explained away given enough data^16, 18^. The remaining aleatoric uncertainty is caused by the variation or noise in the observations, corresponding to the model’s input-dependent (data) uncertainty. Aleatoric uncertainty is *irreducible*, even with more training data^16, 18^.

To capture the model uncertainty^16^, Monte Carlo Drop-Out (MCDO)^14^ performs approximate Bayesian inference via *T* forward passes with random selections of active neurons, given a test case. Similarly, Monte Carlo Batch Normalization (MCBN)^39^ exploits the stochasticity of minibatch statistics in order to obtain predictive distributions. *T* forward passes with random minibatches sampled from training data during inference leads to a predictive distribution for a test case. In this respect, MCBN also captures model uncertainty^39^. However, MCBN explicitly trades the well-informed moving averages of population statistics with those obtained from individual minibatches for the sake of stochasticity in predictions. This leads to a less than optimal discrimination performance of the network^35^, especially given the whole purpose of *Batch Renormalization (BReN)*^36^ that makes a deliberate use of the moving averages and improves on BN. Also, MCBN requires the training data to be available for sampling during inference. The procedure further requires that the same minibatch size be specified during training and inference; otherwise, the respective approximate posteriors would be inconsistent^39^. If a pretrained network is used for inference, it is virtually impossible to figure out the training minibatch size, unless it is documented.

Considering the epistemic and aleatoric uncertainties as properties of the model and the data, respectively, a Bayesian deep learning framework that jointly models both types of uncertainties has been recently proposed^16^. While simultaneous modeling of both enables the best uncertainty calibration, the overall calibration quality has been found to be dominated by the explanation of aleatoric uncertainty. The contribution from the epistemic component is marginal; hence, in the regime of big data, it is more effective to tackle aleatoric uncertainty^16^.

Recently, a non-Bayesian *ensemble* approach has been combined with *adversarial* training^17^. Basically, the ensemble consists of multiple neural networks and it is diversified by their random initialization and random shuffling of training examples. It readily provides predictive distributions and uncertainty estimates during inference. Adversarial examples, which are essentially augmented training examples^17, 69^, explore the local neighborhood of the original training examples to test the robustness of neural networks. As a result, the likelihood of the data smooths out in the *ε*-neighborhood of examples and the ensemble generates well-calibrated outputs^17^, thanks to the capturing of the both uncertainty types together. Ensembles typically work better than MCDO and are robust to dataset shift^70^. Nevertheless, the input-space exploration is guided by gradients and the adversarial examples are attracted towards the objective function^42^. While the procedure avoids the computational burden of the ideal exploration in all directions^17^, it leaves some areas of the input space underexplored^42^. By TTAUG, we aim at efficiently exploring the neighborhood of examples in all directions in order to obtain predictive uncertainty estimates. The median of the TTAUG-distribution of predictive probabilities readily offers well-calibrated outputs and uncertainty estimates (Fig. 5d).

Selective prediction has been recently discussed in the context of DNNs^66–68^. The proposed framework constructs a well-calibrated classifier, given an uncertainty estimation function for the classifier. While the initial selective classifiers adopted pretrained networks and learned selection functions separately^67^, *SelectiveNet*^68^ trains a classifier and its selection function simultaneously. Thus, the classifier focuses on the most relevant examples and offers risk guarantees for its predictions at a given threshold for its coverage of data^67, 68^. So far, this framework has been shown to work with uncertainty estimates from softmax outputs and MCDO^67, 68^. Evaluating TTAUG-based uncertainty estimates in this framework is an important future direction.

### Uncertainty in Medical Image Analysis via DNNs

Even though several strategies have been proposed for quantifying different kinds of uncertainty, their use in medical context is only now gaining traction. For instance, Leibig et al.^28^ used a VGG^71^-like CNN equipped with MCDO for DR detection from fundus images. Leveraging the uncertainty information from their network for decision referral, they obtained improvements in automated DR detection performance. However, we achieved better ROC-AUC scores, given the same disease detection tasks and data, via our TTAUG-based uncertainty measure. Moreover, in a recent study of brain tumor segmentation^53^, uncertainty estimates via data augmentation not only helped improve the segmentation accuracy but also outperformed the epistemic uncertainty captured by MCDO in segmentation quality.

### Outlook

This study demonstrates the effectiveness of TTAUG in capturing the predictive uncertainty of DNNs and its concordance with the uncertainty in clinicians’ decisions on a well-known diagnostic task. However, the procedure only works on the outputs of a network which itself is not capable of exploiting the estimated uncertainty. By assuming the predictive uncertainty of DNNs as a proxy for interobserver variability and integrating it into training, we may emulate the transfer of human grader variability to DNNs and facilitate the cooperation of man and machine in the form of assisted reading^23^. The selective classification framework^67, 68^ may provide a starting point for this. This approach could be extended to visualization^9, 72–75^ efforts in the hope of improving the interpretability of DNN decisions in medical settings.

## Data Availability

All data used in this study are publicly available. More information can be found via the following links:
Dataset 1: https://www.kaggle.com/c/diabetic-retinopathy-detection
Dataset 2: https://idrid.grand-challenge.org/

## Acknowledgements

We thank Christian Leibig for many useful discussions, helpful suggestions as well as providing the preprocessed Kaggle DR images. This research was supported by the German Ministry of Science and Education (BMBF, 01GQ1601 and 01IS18039A) and the German Science Foundation (BE5601/4-1 and EXC 2064, project number 390727645).

## Author contributions statement

MSA and PB designed research; MSA performed research; LK, GA, WI and FZ provided medical advice and graded images; PB supervised research; MSA and PB wrote the paper with input from all authors.

## Footnotes

1 Minibatches of 23 consists of 15,2,4,1 and 1 examples from each class, which still induces an oversampling of the minority classes; however, it preserves the majority of the No DR examples. Also note that 23 is the maximum number of examples that fit into the GPU memory per iteration.

